# Prevalence of Polycystic Ovary Syndrome and Its Impact on Psychological Distress: A Comparative Study among Urban and Rural Reproductive-aged women in Bangladesh

**DOI:** 10.1101/2025.07.03.25330821

**Authors:** Anup Talukder, Tahmina Akter Tithi, Maruf Hasan Rumi, Abdul Muyeed

## Abstract

Polycystic ovary syndrome (PCOS) is a common hormonal disorder of reproductive-aged women all over the world. Low- and middle-income countries, such as Bangladesh, pay less attention to mental health issues. As a result, the prevalence of psychological issues like depression, anxiety, stress, and insomnia, is increasing alarmingly, as PCOS has a negative impact on these detrimental aspects. The study assesses the prevalence and psychological impacts of PCOS among reproductive-aged women as well as compared the mental health inequalities between rural and urban women in Bangladesh. A sample of 212 reproductive-aged women was collected using a convenience sampling procedure. Additionally, for diagnosis of PCOS, the Rotterdam criteria were used. Depression, anxiety, stress, and insomnia were measured using DASS-21 and ISI-7 tool. To analyse the association and impact of PCOS on mental health problems, a chi-square test and multivariate binary logistic regression were conducted. The Mann-Whitney U test was implemented to assess mental health disparities among rural-urban reproductive women. Among the participants, the PCOS prevalence was 50.9%, and the rates of depression, anxiety, stress, and insomnia were 51.4%, 62.3%, 46.7%, and 53.3%, respectively. The prevalence’s were significantly greater in women with PCOS than those without PCOS. Women residing in rural region were more prone to meet Rotterdam criteria and had higher anxiety and insomnia syndrome. Women who met two or three diagnostic criteria had a higher chance of experiencing psychological distress than others. Furthermore, relationship status and BMI are useful predictive factors of mental health consequences. The findings showed a substantial prevalence of emotional distress among PCOS women in Bangladesh. Additionally, a comprehensive mental health investigation is suggested for PCOS. Addressing both the physical and psychological aspects of PCOS is essential to develop women’s overall well-being and quality of life.

**Highlights:**

- Women with PCOS showed a higher prevalence of emotional distress.
- Women meeting two or three Rotterdam criteria had higher prevalence of distress.
- Rural women had higher risk of having anxiety and insomnia.
- Women who engaged in a relationship had higher chance of having depression.
- Overweight women had higher risk of experiencing depression.

## 1.0 Introduction

Polycystic ovary syndrome (PCOS) is one of the most prevalent endocrine disorders among reproductive-aged women worldwide [1]. This disorder produces extreme amount of androgen from the ovaries because of its multidimensional and polygenic behavior. It also comprises dysregulation of CYP11a gene and upregulation of the enzymes involved in androgen biosynthesis [2]. Furthermore, it is also linked to the insulin receptor gene, located on the chromosome 19p13. 2 [3]. According to the World Health Organization, it affects between 6 and 10% of reproductive-age women and an estimated 70% of cases are undiagnosed worldwide. This syndrome has large influence on the female biological aspect such as obesity, body image and infertility, leading to social stigma. It is also having a negative contribution in mental health such as psychological well-being, depression, anxiety, stress, insomnia among women who are in reproductive age across the world [4]. The worldwide prevalence of PCOS is estimated to be 9.2% [5]. A prevalence of 11.4% has been reported in the south Asian region [6]. However, the incidence of PCOS and subsequent mental health disorders in South Asian women appears to be substantially higher than the worldwide average. This is due to some reasons such as social and cultural taboo, religious and political diversity, collectivist society, and a shortage of mental health services [7–9]. Moreover, PCOS women were more likely to contribute certain physical syndromes, include menstrual irregularity or infertility, obesity, insulin resistance, dyslipidaemia, hypertension and metabolic dysfunction rather than PCOS free, which may subsequently contribute to cardio metabolic disorders [10]. Despite the physical issues, it has also significant association with some psychological illnesses such as depression, anxiety, stress, insomnia, bipolar disorder, obsessive compulsive disorder, and somatization disorders [11,12]. Other frequently reported symptoms include concentration difficulty, low self-esteem, loss of interest or pleasure, fatigability, increased fatigue, reduced activity, appetite decrease and possible suicidal ideation [13]. The relationship between mental health condition and PCOS implies that increase in prevalence of PCOS may worsen the circumstance of mental health.

In South and Southeast Asian nations, including India and Thailand, studies have shown that women with PCOS endure comparable psychological distress, often exacerbated by insufficient healthcare access and societal stigmas. Keeratibharat et al. (2024) conducted a study revealing that the prevalence of depression, anxiety, and poor mental well-being among Thai women with PCOS was 3.8%, 11.9%, and 16.9%, respectively [14]. Studies in India have shown that women with PCOS have high levels of anxiety and depression because they can’t get the treatment they need [9].

Nonetheless, owing to a lack of resources and information, developing nations like Bangladesh are confronting the challenge of mental health burdens [15]. Hasan et al. (2024) executed a cross-sectional study involving 409 PCOS patients, revealing prevalence rates of loneliness, anxiety, and depressive syndrome at 71%, 88%, and 60%, respectively [16]. Additionally, Ishrat et al. (2021) recruited 126 consecutive infertile women with PCOS for a cross-sectional study, demonstrating that obesity correlates with hyperandrogenism, hyperinsulinemia, insulin resistance, and metabolic syndrome [17]. Furthermore, a cross-sectional study by Kamrul-Hasan et al. (2021) on adolescents with PCOS revealed that an elevated BMI heightens the risk of metabolic syndrome [18]. Sarker et al. (2015) reported in their study that the greatest clinical predictor for PCOS diagnosis in infertile women is the presence of hirsutism [19]. Numerous cultural and societal variables exacerbate the problems associated with PCOS in low- and middle-income nations like Bangladesh [20,21]. Consequently, a large number of women experience numerous chronic mental health problems as a result of being undetected and untreated. Despite the fact that a great deal of research has been done globally on the psychological implications of PCOS, Bangladesh has gotten less attention.

Existing research has investigated the impact of PCOS on mental health focusing on depressive syndrome, anxiety, loneliness as well as its clinical, hormonal and metabolic parameters. However, few researches have thoroughly examined the psychological problems that PCOS patients in Bangladesh face, such as stress and insomnia. Therefore, the study’s primary goal is to assess the prevalence of emotional distress (depression, anxiety, stress, and insomnia) and polycystic ovarian syndrome (PCOS) among Bangladeshi reproductive aged women while also examining the effects of PCOS on emotional distress. It also examines the group differences in depression, anxiety, stress, and insomnia amongst Bangladeshi women living in urban and rural areas. The study aims to clarify the psychological burden faced by women with PCOS and reinforce the necessity for comprehensive treatment by examining the prevalence and possible causes of these issues.

## 2.0 Materials and Method

### 2.1 Study design

A quantitative and cross-sectional methodology was used to conduct the study, which involved women in Bangladesh who were of reproductive age between March 1, 2025, and April 15, 2025. Hasan et al. (2024) and Kite et al. (2021) utilized comparable methodologies in their investigations on associated subjects [16,22]. Data was collected from respondents via a face-to-face interview using a structured questionnaire that comprised four distinct sections.

### 2.2 Study participants and Sample

Women in the reproductive age range of 15–49 years were enrolled in the study [23], regardless of whether they had PCOS or not. The sample size was calculated using Godden’s algorithm with a 5% margin of error and a 5% threshold of significance (α = 0.05). A cross-sectional survey that found an 11.92% prevalence of anxiety among Thai women with PCOS was used to calculate the study’s sample size [14].

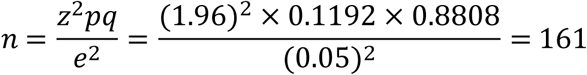

It was determined that 161 was the ideal sample size. 220 reproductive female participants, ages 15 to 49, were given the questionnaire; regardless of whether they had PCOS or not, all of them acknowledged the need for an ultrasound screening. After excluding eight people with metabolic abnormalities, hormonal medication, acute psychiatric concerns, and incomplete data, 212 participants were ultimately chosen for the study with their consent. Out of the 212 respondents who had been selected, 108 were diagnosed with PCOS after meeting at least two of the Rotterdam criteria. However, since they satisfied fewer than two diagnostic criteria, the other 104 respondents were categorized as not having PCOS. In Fig 1, a sampling flowchart was displayed.

**Fig 1.**
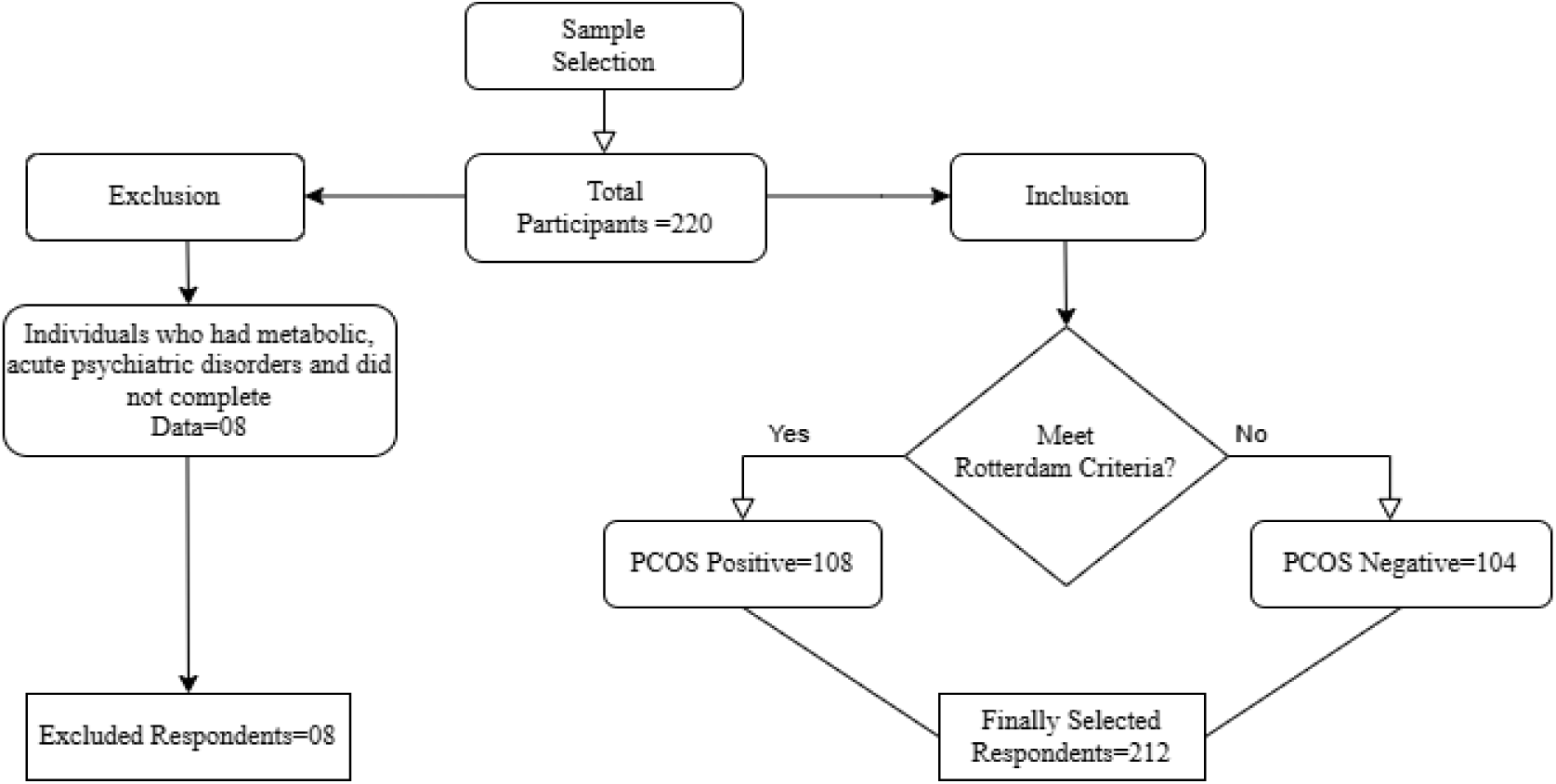
Sampling Flow chart with study inclusion and exclusion criteria

### 2.3 Instrument and data collection

The participants were chosen using a convenience sampling method. In Bangladesh, reproductive health conditions are almost the same in every administrative division [24]. For convenience, the study was carried out in Mymensingh with the goal of cutting down on research expenses and time. The study’s area map was shown in Fig 2. Data were collected from four government hospitals located in four administrative districts within the Mymensingh division. The institutions comprise Mymensingh Medical College and Hospital, Netrokona Medical College and Hospital, Jamalpur Sadar Hospital, and Sherpur Sadar Hospital. A total of 150 data had been gathered by collecting 50 responses from each of three hospitals: Netrokona Medical College and Hospital, Jamalpur Sadar Hospital, and Sherpur Sadar Hospital. Following that, 62 data were obtained from the remaining hospital, Mymensingh Medical College and Hospital. The data collectors collected the relevant data for the study from reproductive-aged women exiting the gynecology departments who met the inclusion criteria and gave consent Data was gathered by conducting an in-person interview and recording the answers with a structured questionnaire. Participants received printed versions of the questionnaires. A validated four-section questionnaire was created in Bengali and English. To make sure the survey was valid and reliable; a pilot study with ten respondents was carried out. The Rotterdam criteria, which are the diagnostic criteria for PCOS, are incorporated into the questionnaire in the first section. Hyperandrogenism, irregular menstruation, and polycystic ovaries on ultrasound are the three Rotterdam criteria for diagnosing PCOS. At least two of these three criteria had to be met in order to diagnose PCOS. Background information was provided in the second section. The Depression, Anxiety, and Stress Scale (DASS-21) was used in the third segment, and the Insomnia Severity Index (ISI-7) was used in the last.

**Fig 2.**
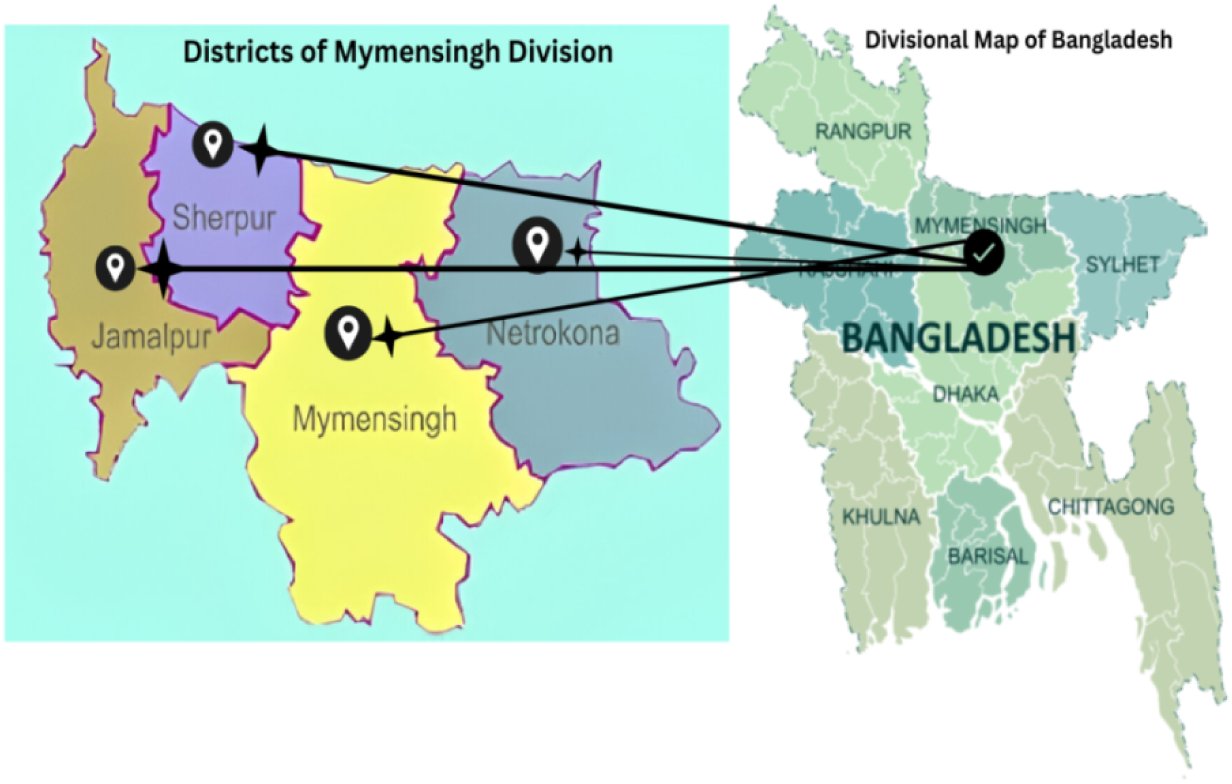
Map of the study area [25]

### 2.4 Depression, Anxiety, Stress Scale-21 (DASS-21)

Depression, Anxiety, and Stress Scale (DASS-21) is used to assess participants’ overall psychological well-being over the past few weeks. The 21 questions on the DASS-21 are composed of seven items from the subscales for stress, anxiety, and depression. A 4-point Likert scale with a range of 0 to 3 is used with a sequence of statements to show the degree of agreement. The following is the definition of the rating scale: The numbers 0 through 3 represent not at all, some of the time, good part of the time, and most of the time, respectively. The relevant category scores are summed together, and the overall score is then multiplied by two to approximate the degrees of stress, anxiety, and depression. A normal level of anxiety is indicated by a total score between 0 and 7. Furthermore, mild, moderate, severe, and extremely severe anxiety are represented by the score ranges of 8–9, 10–14, 15–19, and 20+, respectively. Four different levels are identified in the total score range for stress: (0–14), (15–18), (19–25), (26-33), and 34+. These levels represent normal, mild, moderate, severe, and extremely severe stress, respectively [26]. A detrimental state for stress, anxiety, and depression is indicated by scores that range from moderate to extremely severe. In order to ensure the reliability and validity of the translation, a faculty member from Jatiya Kabi Kazi Nazrul Islam University’s English department verified the Bengali translation of the original DASS-21 scale as part of the validation procedure. In this case, it was determined that Cronbach’s *⍺* coefficient of this scale was 0.920, suggesting that it exhibits high internal consistency and reliability.

### 2.5 Insomnia Severity Index (ISI-7)

A sleep disorder called insomnia makes it difficult to fall asleep. Additionally, chronic insomnia has been linked to a number of major health concerns, such as metabolic syndrome, type 2 diabetes mellitus, cardiovascular disease, weight-related difficulties, hypertension, dyslipidemia, and colorectal cancer [27]. The insomnia severity index is a widely used measurement tool to assess the degree of sleep-related problems over the previous two weeks. Seven items with a five-point Likert scale comprise the evaluation. The insomnia total score ranges from 0 to 28. Insomnia is more severe when the score is more than 14. Chronic insomnia is indicated by a higher total insomnia score. The sum of the scores from all seven questions is used to determine how severe insomnia is. The following is an explanation of how the overall scores were categorized: Clinically severe insomnia is absent when the overall score falls between 0 and 7. The overall scores fall into three categories: “sub-threshold”, “moderate”, and “severe clinical insomnia” [28]. These are 8–14, 15–21, and 22–28, respectively. With a Cronbach’s *⍺* value of 0.923, the scale was found to have a good degree of internal consistency and reliability. A professor from Jatiya Kabi Kazi Nazrul Islam University’s English department verified the Bengali translation of the original ISI scale as part of the validation procedure.

### 2.6 Socio-demographic characteristic

Participants were asked about their demographics and socioeconomic status, and it was assured that their data would be kept private. The factors taken into account were socio-demographic factors: age, height, weight, parity, smoking and drinking habits, physical disabilities, other diseases, family environment, residential location, relationship status, educational background, and occupation.

### 2.7 Statistical Analysis

The research employed descriptive statistics to illustrate categorical variables, emphasizing frequency and percentage. The Depression, Anxiety, Stress Scale-21 (DASS-21) and the Insomnia Severity Index-7 (ISI-7) were utilized to measure the severity of mental health issues, including depression, anxiety, and stress, as well as insomnia. Additionally, Cronbach’s *⍺* coefficient of internal consistency was utilized to assess the reliability of DASS-21 and ISI-7 [29]. The ideal cut-off for Cronbach’s *⍺* is 0.70, with the suggested range extending from zero to one. A value approaching 1 and exceeding 0.70 signifies improved internal consistency or reliability [30]. To examine the association between PCOS, emotional distress, and socio-demographic characteristics, a bivariate chi-square analysis was performed. The influence of PCOS on mental health problems and insomnia was then to be clarified by examining the cause-and-effect relationship of significant related factors using multivariate binary logistic regressions (MBLR) with a 95% confidence interval. Furthermore, the Mann-Whitney U test was used to evaluate the disparities in depression, anxiety, stress, and insomnia between Bangladeshi women residing in urban and rural areas. Before conducting the Mann-Whitney test, the assumption of distribution symmetry between groups was checked using QQ plots. As a result, the finding ensured that the distribution was similarly shaped that validated the use of Mann Whitney test for comparing median. All forms of statistical analysis were carried out using IBM Incorporation’s Statistical Package for Social Science (SPSS, version 25). Additionally, R programming was used to create the prevalence graph.

### 2.8 Ethical clearance

Ethical approval for this study was granted by the Research Ethics Committee of the Public Health Foundation Bangladesh (PHFBD) in Dhaka. The study adhered to the ethical principles outlined in the Declaration of Helsinki. Written informed consent was obtained from all participants prior to data collection. The research protocol was approved under reference number PHFBD-ERC-NFP-E-18/2025.

At the beginning of data collection, participants received a thorough explanation of the study’s objectives, aims, methods, affiliations, benefits, and hazards at the planned face to face interview session. Afterward, the respondents agreed to take part in the study, and the data collectors promised that their answers would be kept private and not shared. With their consent, data was subsequently gathered from them for the research purpose.

## 3.0 Results

Table 1 depicts factor-wise PCOS prevalence and associations between socio-demographic variables and depression, anxiety, stress, and insomnia. Most respondents are students (81.6%) with honors-level education (63.2%). A majority of them are single (56.6%) and live in urban areas (53.3%). On the other hand, BMI is equally divided between underweight and normal weight categories (33% each). Moreover, Fig. 2 represents the prevalence of PCOS and mental health condition. Among the respondents, more participants have PCOS present (50.9%) than absent (49.1%), while depression (51.4%), anxiety (62.3%), stress (46.7%), and insomnia (53.3%) are prevalent. Based on the residential areas of the respondents, rural women had higher levels of anxiety (p<0.001) and stress (p<0.05), while urban women experienced lower insomnia (p<0.01). Furthermore, depression (p < 0.001) and anxiety (p<0.05) were significantly associated with obesity (BMI ≥30). On the other hand, women with PCOS present had significantly higher levels of depression, anxiety, stress and insomnia compared to those absent (p<0.001). Additionally, women meeting two or three Rotterdam criteria exhibited significantly higher rates of depression, anxiety, stress, and insomnia (p<0.001). Fig 3. presents the prevalence of PCOS and mental health condition among reproductive-aged women in Bangladesh.

**Fig 3:**
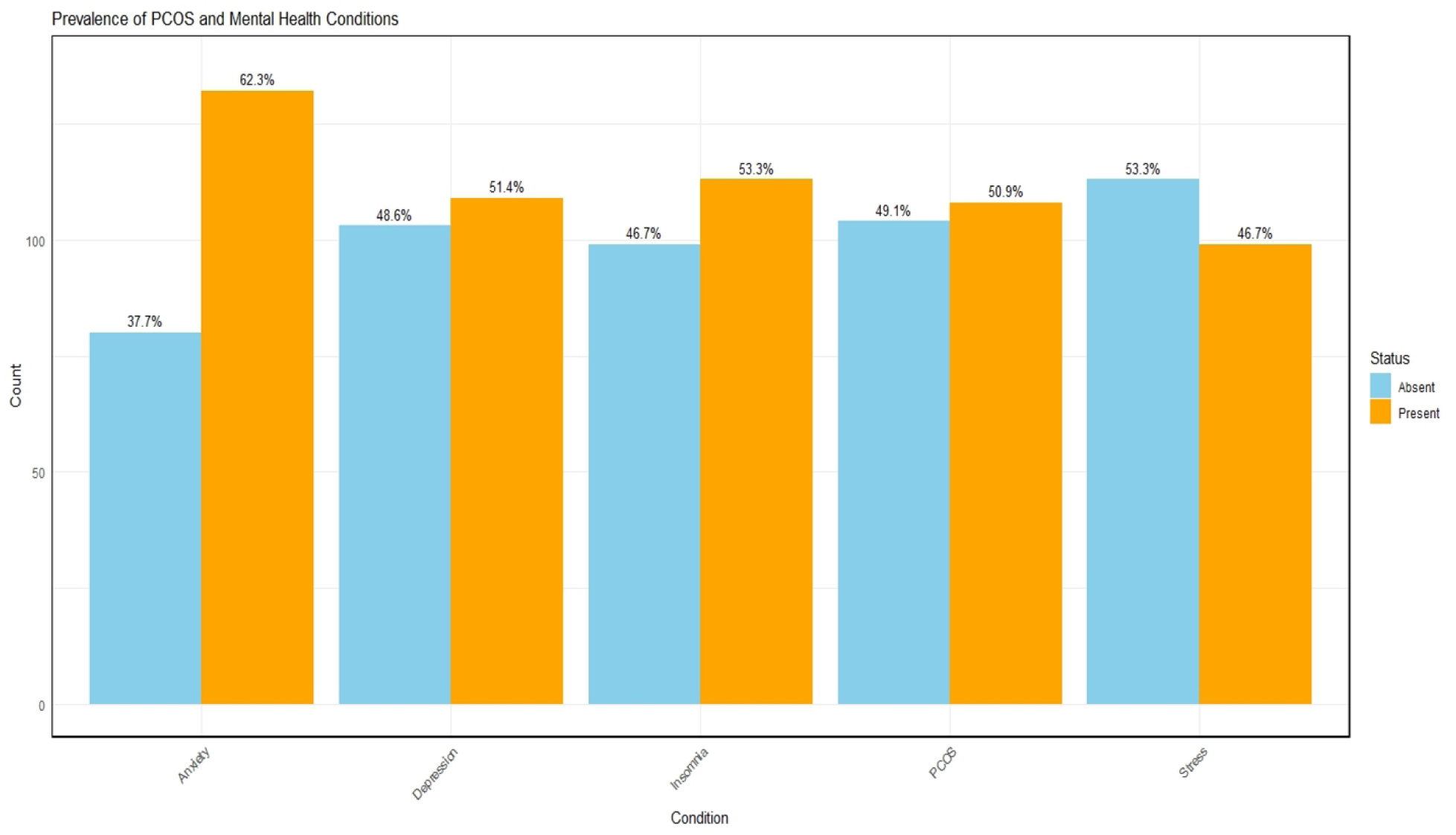
Prevalence of PCOS and Mental Health Condition among Reproductive-aged Women in Bangladesh.

**Table 1.**
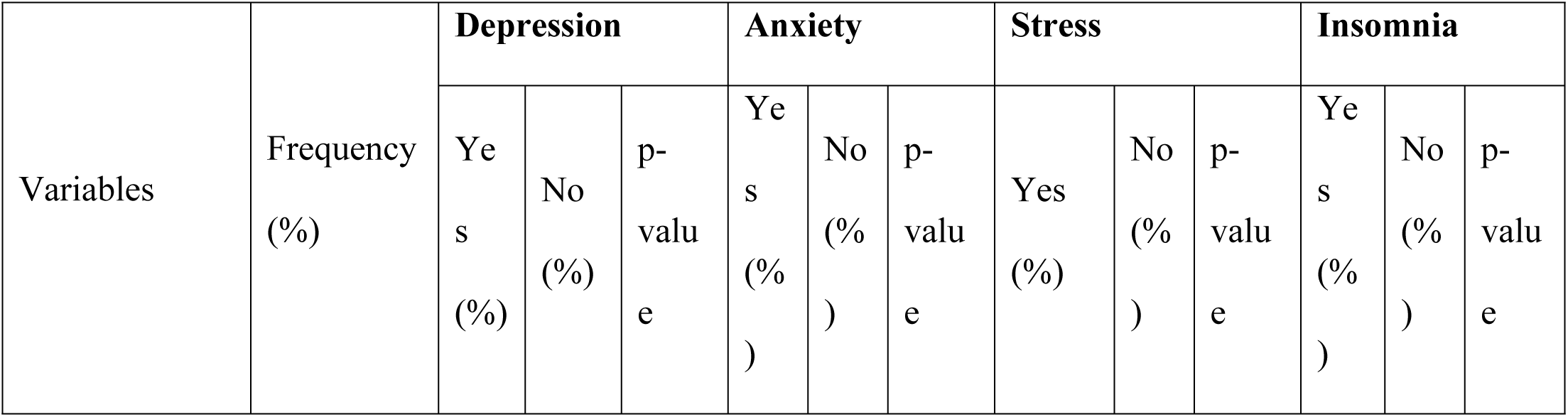
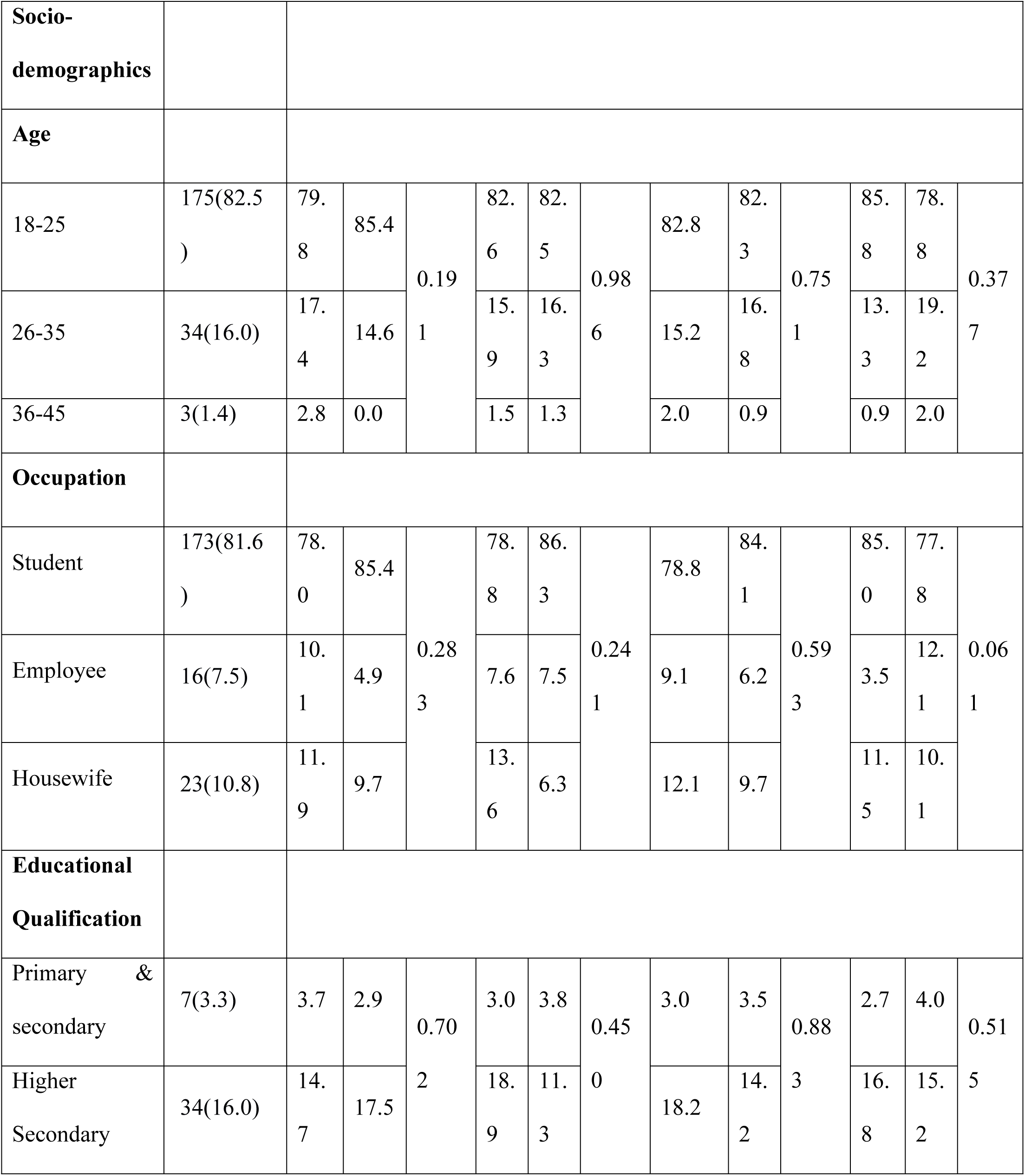
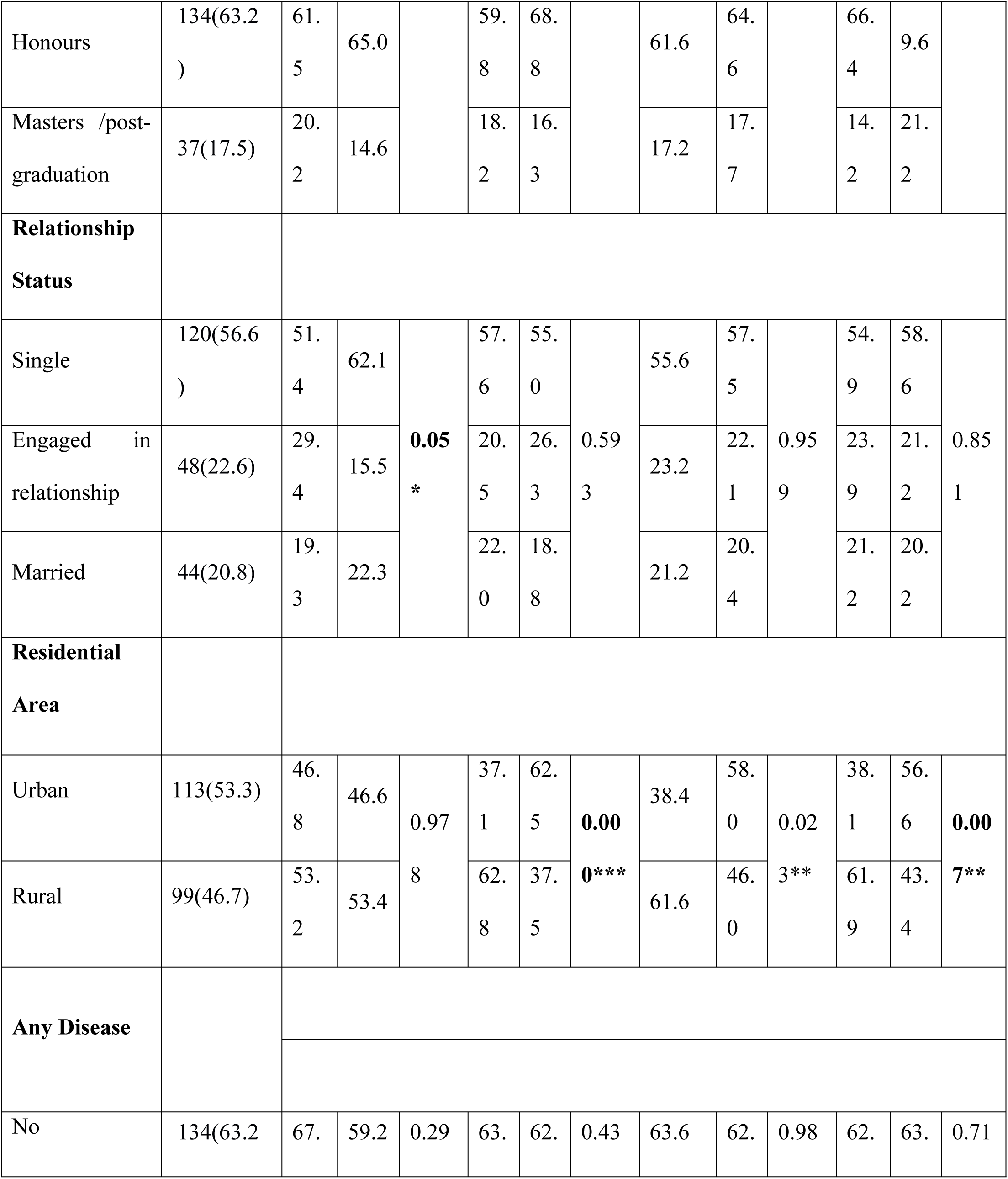
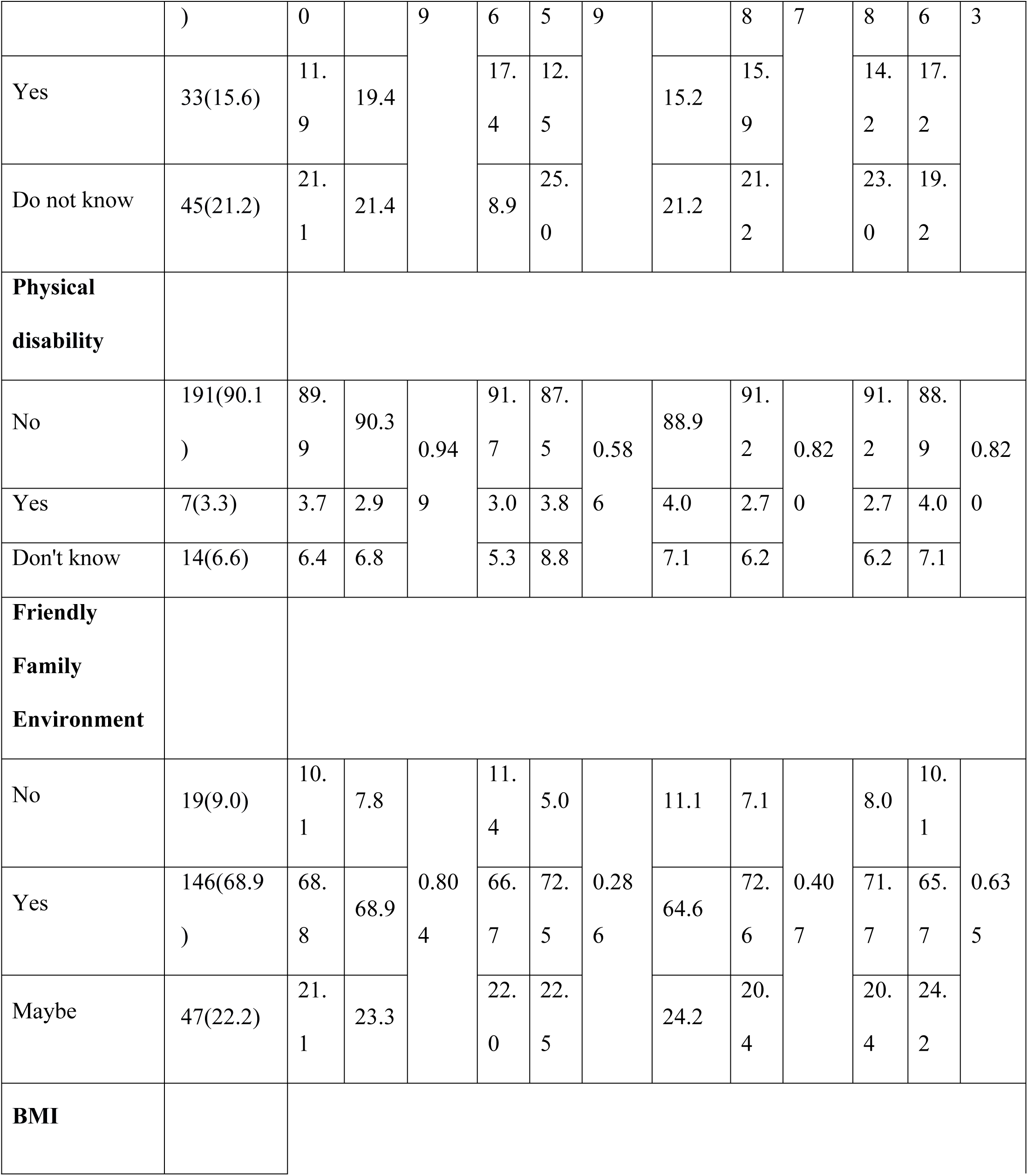
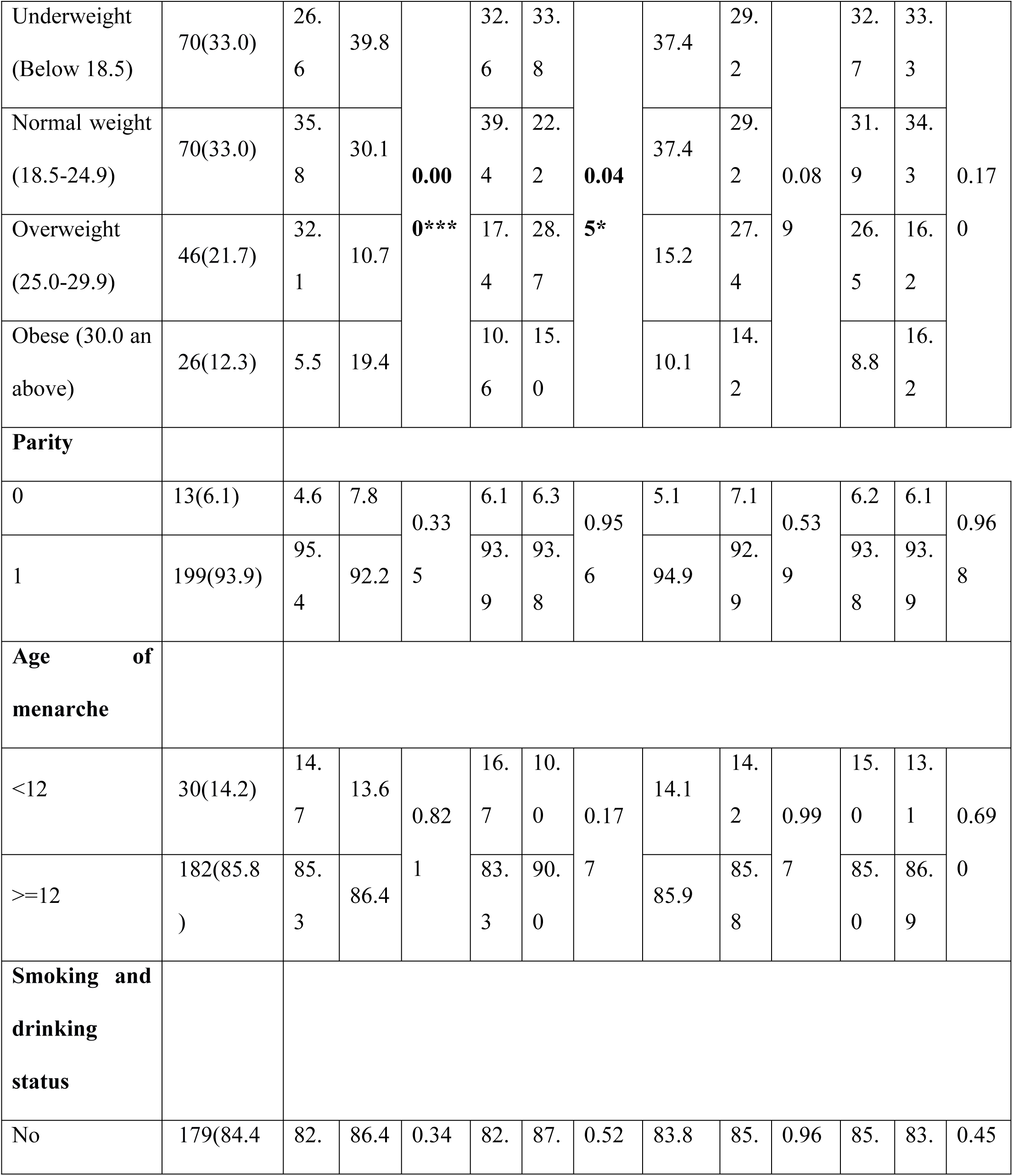
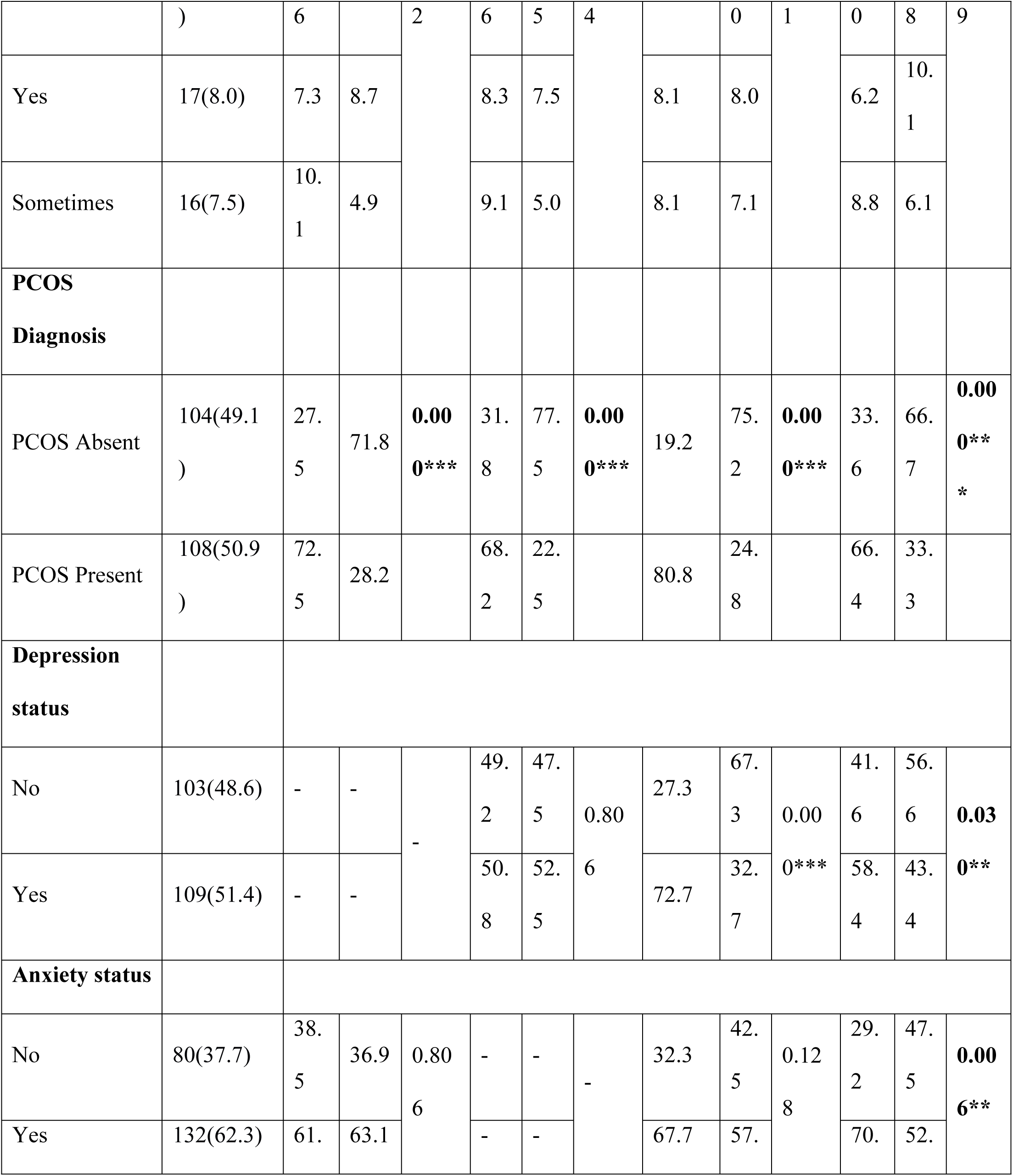
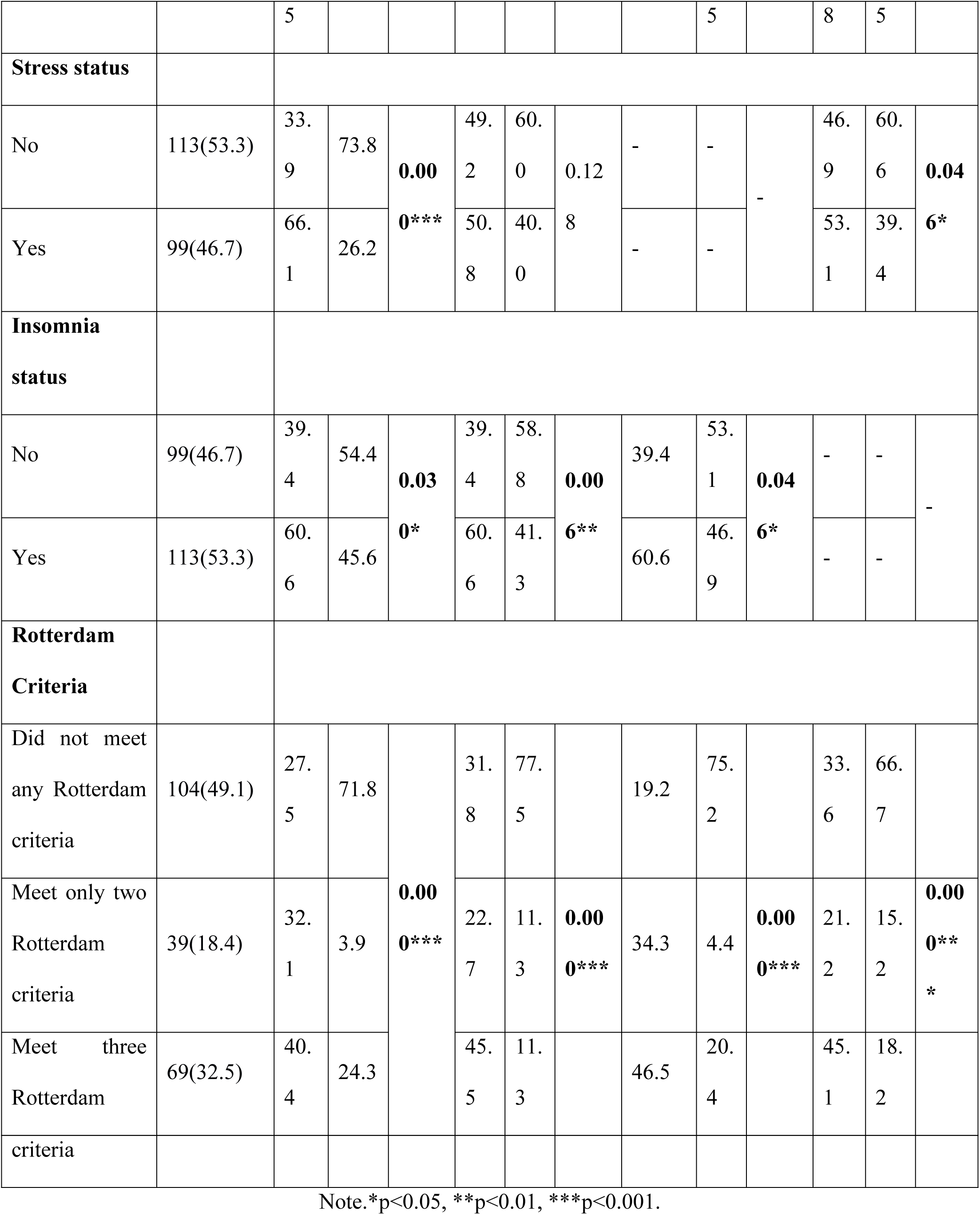
Factor-wise PCOS prevalence and associations between socio-demographic variables and depression, anxiety, stress, and insomnia.

Table 2 illustrates the binary logistic regression analysis for factors associated with depression, anxiety, stress, and insomnia. According to the table, being engaged in a relationship increases the likelihood of depression (aOR=2.676) with a significant association (p<0.05). In addition, normal weight increases the risk for depression (aOR=2.450, p<0.05) and anxiety (aOR=2.601; p<0.05), while overweight individuals are more likely to experience depression significantly (aOR=8.802, p<0.001). On the other hand, such mental health issues are associated with having PCOS (all conditions, p<0.001). However, meeting two or three Rotterdam criteria is significantly associated with all four mental health issues.

**Table 2.**
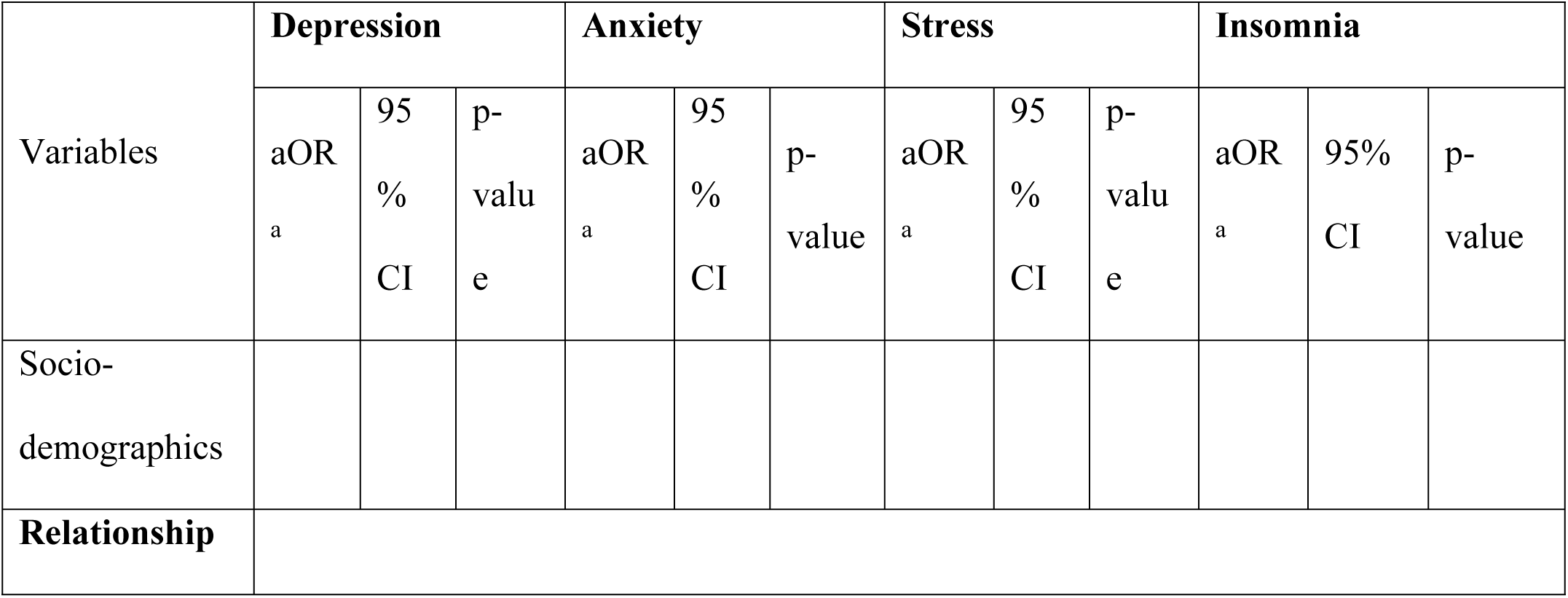
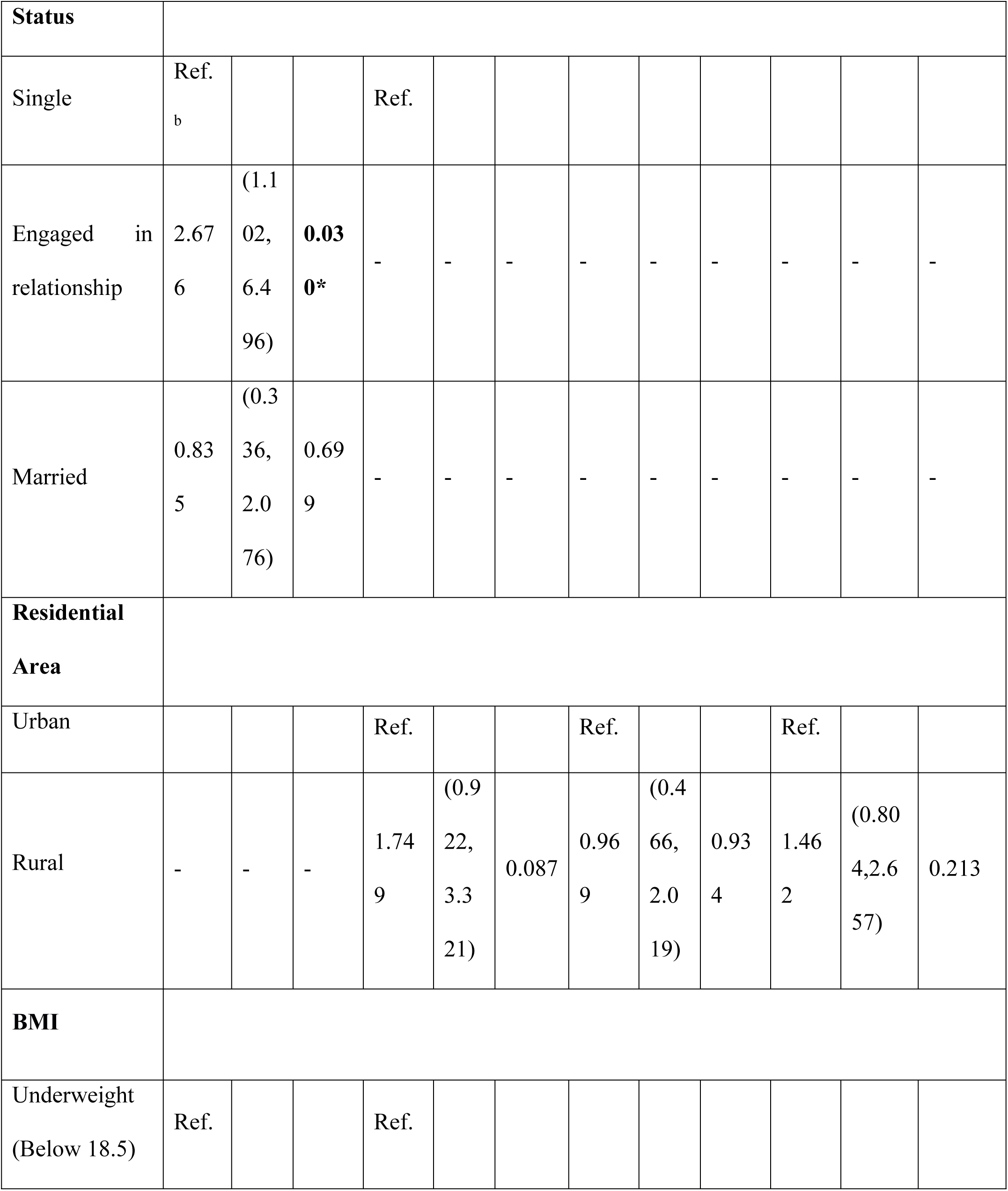
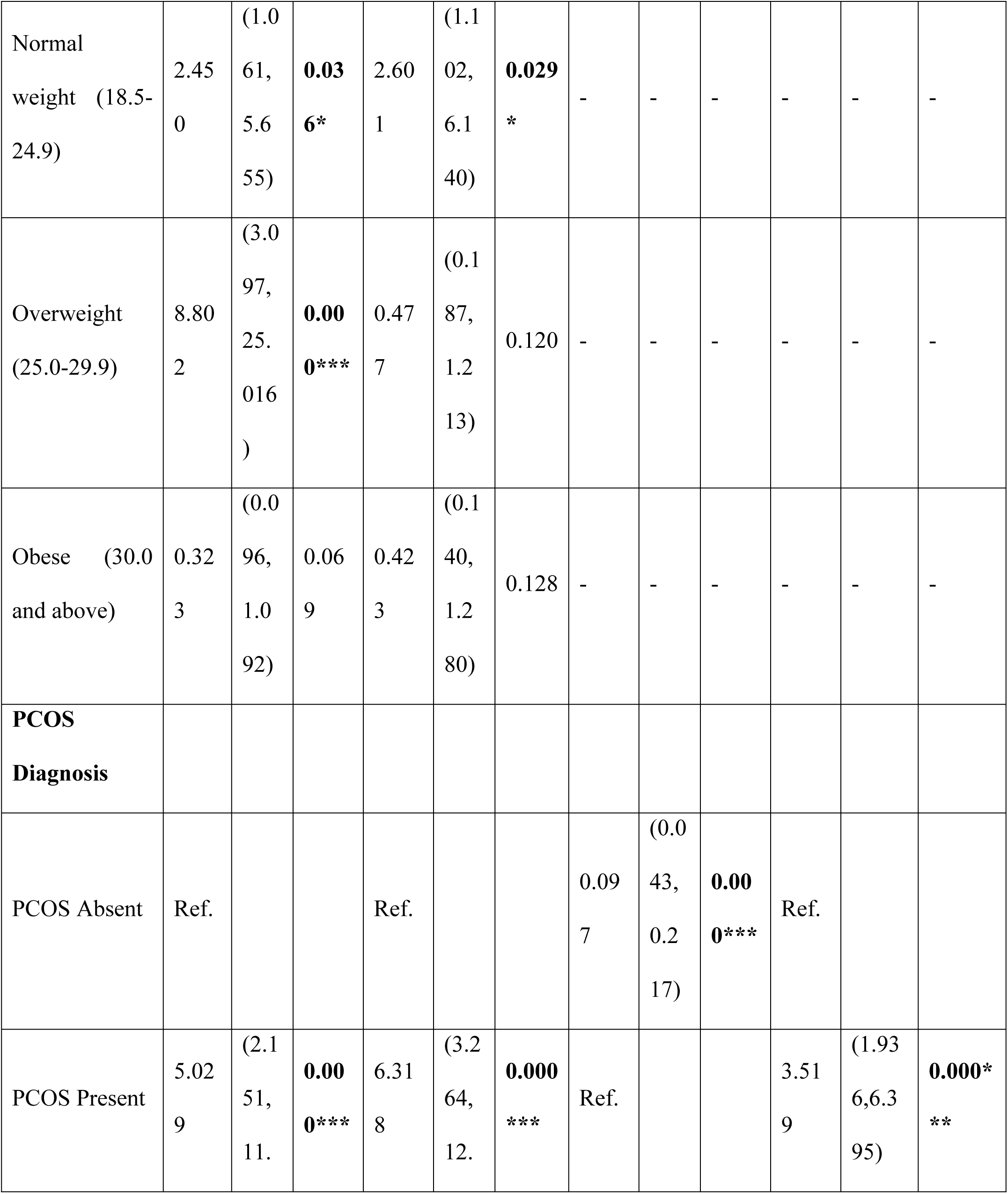
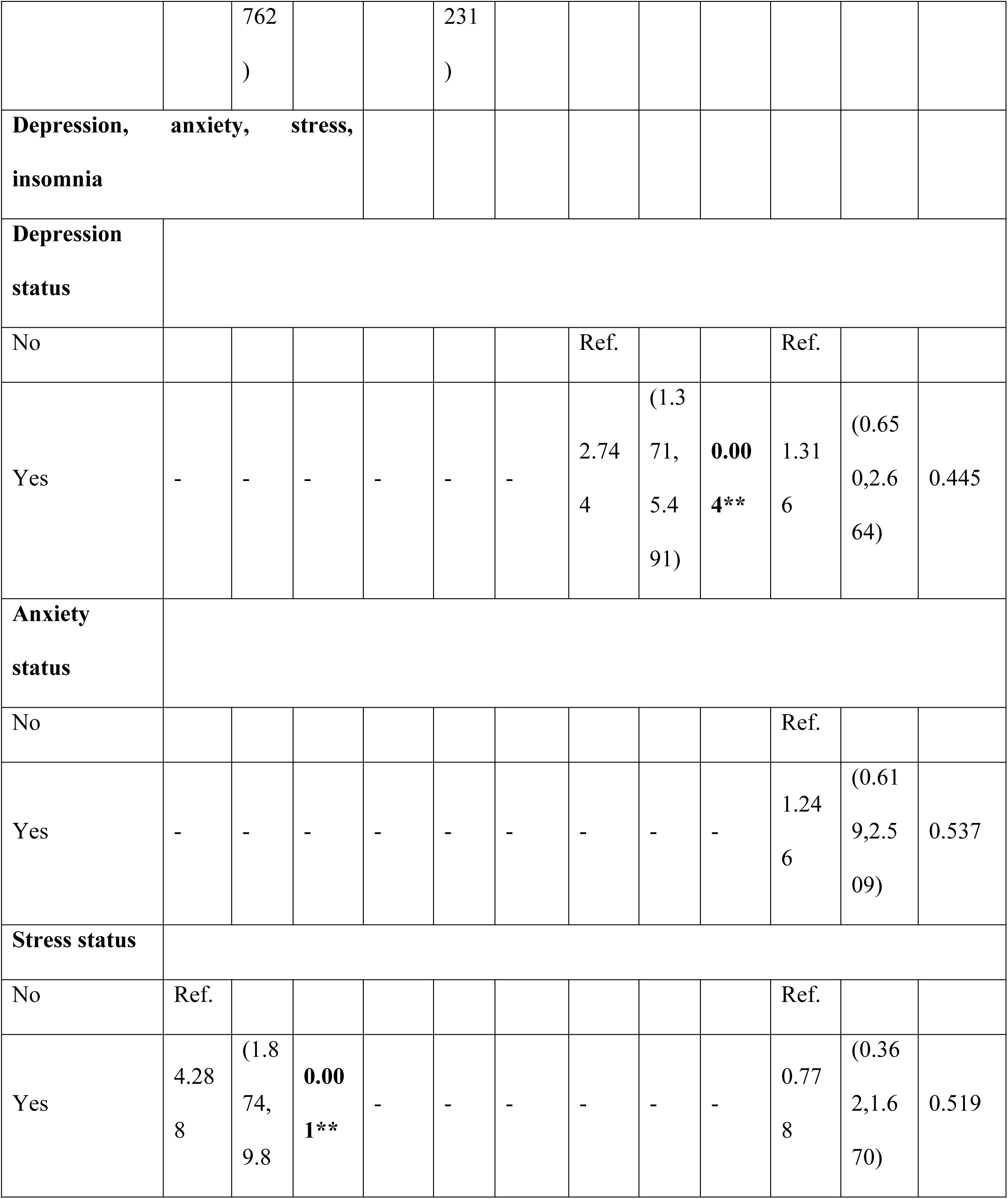
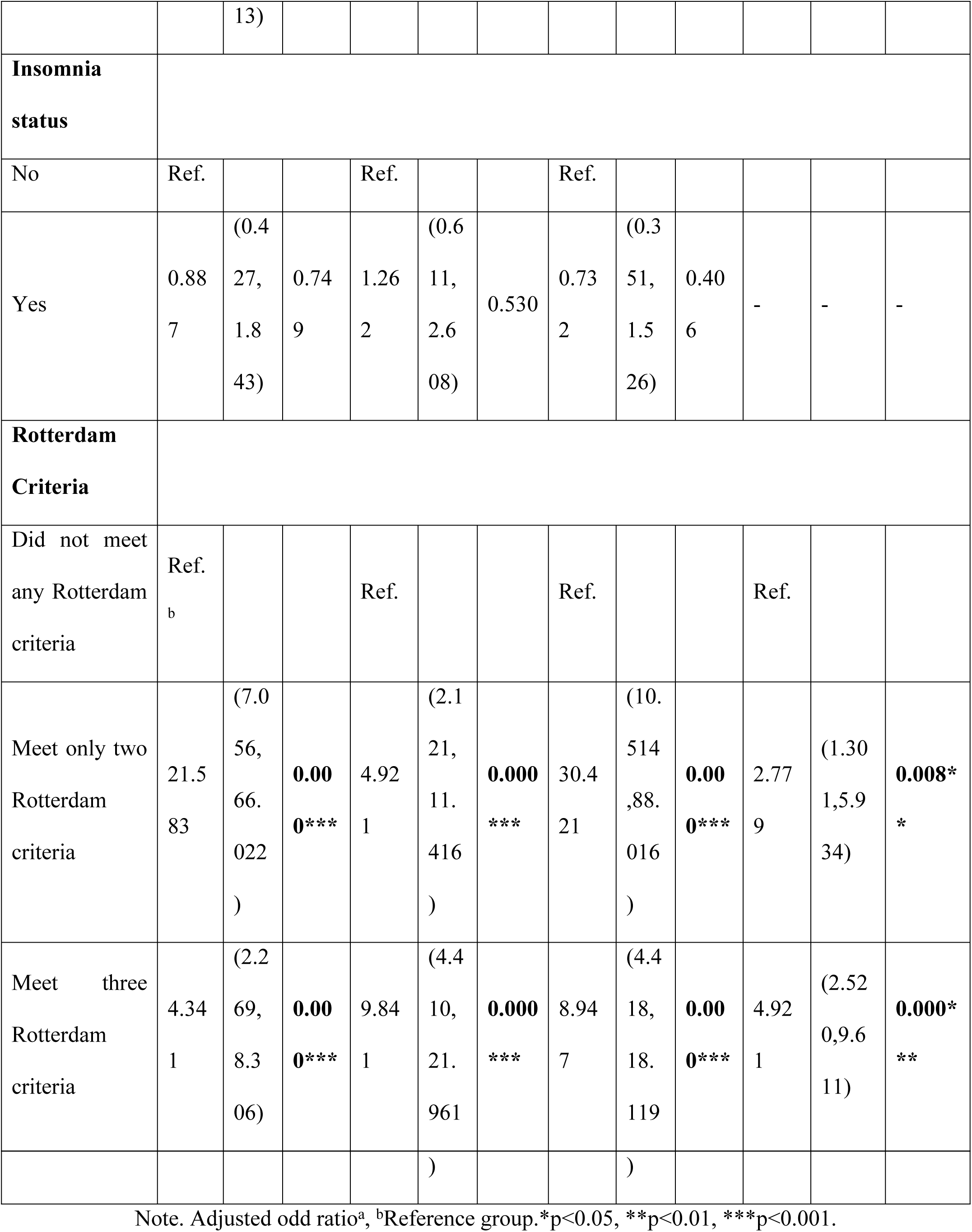
Binary Logistic Regression Analysis of Factors Associated with Emotional Distress.

Table 3 demonstrates the association of Rotterdam criteria across residential areas. The table shows significant differences in the association of Rotterdam criteria between urban and rural areas (p<0.001). On the other hand, meeting only two Rotterdam criteria had 2.5 times higher odds (aOR = 2.497) of meeting the criteria compared to those who did not, with a significant (p<0.05) in rural areas. Additionally, meeting all three Rotterdam criteria had 5.3 times higher odds (aOR = 5.313) with a highly significant association (p<0.001).

**Table 3.** Association of Rotterdam Criteria with Residential Area: Chi-Square and Logistic Regression Analysis.

| Chi-square |  |  |  | Binary Logistic Regression |  |  |
| --- | --- | --- | --- | --- | --- | --- |
| Variables | Residential Area |  | P- value | Residential Area |  |  |
|  | Urban (%) | Rural (%) |  | aOR<sup>a</sup> | 95% CI | p-value |
| Rotterdam Criteria |  |  |  |  |  |  |
| Did not meet any Rotterdam | 66.7 | 33.6 | 0.000**<br>* | Ref. |  |  |
| criteria |  |  |  |  |  |  |
| Meet only<br>two<br>Rotterdam<br>criteria | 16.2 | 20.4 |  | 2.497 | (1.176,5.299<br>) | **0.017\*** |
| Meet three<br>Rotterdam<br>criteria | 17.2 | 46.0 |  | 5.313 | (2.698,10.46<br>1) | **0.000\*\*\*** |

Table 4 illustrates group differences in depression, anxiety, stress and insomnia between Urban and rural women diagnosed with PCOS. Considering depression, no significant difference was found (μ=0.749), while anxiety was found with a significant difference (p<0.01) between urban women (μ=14.06) and rural women (μ=17.29). Furthermore, insomnia was more prevalent in rural women (μ=15.01), with a significant difference (p<0.05) which indicates that anxiety and insomnia are significantly higher in rural women with PCOS.

**Table 4.** Group difference in Depression, Anxiety, Stress and Insomnia between Urban and rural Women with PCOS.

| Emotional<br>distress domain | Urban | Rural | Mann-Whitney U-test (Z) | p-value |
| --- | --- | --- | --- | --- |
| *Depression* |  |  |  |  |
| Mean (SD) | 14.59 (5.759) | 14.85 (5.987) | 5452.000 (-.320) | 0.749 |
| Median (IQR) | 16.0 (10) | 14.00 (10) |  |  |
| *Anxiety* |  |  |  |  |
| Mean (SD) | 14.06 (7.513) | 17.29 (7.010) | 4332.500 (-2.870) | **0.004\*\*** |
| Median (IQR) | 8.00 (14) | 20.00 (14) |  |  |
| *Stress* |  |  |  |  |
| Mean (SD) | 17.13 (8.311) | 19.03 (8.435) | 4830.000 (-1.720) | 0.085 |
| Median (IQR) | 14.00 (14) | 22.00 (14) |  |  |
| *Insomnia* |  |  |  |  |
| Mean (SD) | 13.78 (4.741) | 15.01 (5.734) | 4714.000 (-1.979) | **0.048\*** |
| Median (IQR) | 14.00 (6) | 16.00 (8) |  |  |

## 4.0 Discussion

Polycystic ovary syndrome (PCOS) is an extensive and under diagnosed endocrine disorder among women, which have significant impact on their physical and mental health during reproductive ages [31]. In Bangladesh, inadequate healthcare access, low awareness, and cultural burdens surrounding fertility exaggerate the psychological burden of such disorder. According to the study, PCOS prevalence was 50.9%. This result also aligns with a number of South Asian hospital-based studies that used the Rotterdam criteria to report PCOS prevalence rates between 40% and 55%. For instance, the prevalence of PCOS was reported to be 61% and 72.47% in Bangladesh and India respectively [32,33]. Moreover, societal stigma, gender norms, and rural-urban differences further deteriorate emotional distress among affected women [34].

The findings of this study showed that women diagnosed with PCOS were around 10 times more likely to experience emotional distress compared to women without the disorder. This estimation surpasses previous studies that reported an 8.32-fold intensification in the probability of stress among PCOS patients [35]. Moreover, women experiencing stress had a 4.2-times higher likelihood of developing depression, underpinning the well-established comorbidity between stress and depression in such women. Thus, these findings are consistent with the stress theory, which posits that chronic physical conditions like PCOS works as persistent stressors that intensify psychological vulnerability, predominantly to anxiety and mood disorders [36].

In addition, sleep disturbance is another prominent mental concern that associated with diagnosed PCOS. Findings showed that 53.3% of women with PCOS experienced insomnia that is a much higher rate than previously reported among Polish (10.5%) and UK (44.2%) women with PCOS [22,37]. This heightened prevalence reinforced the stress process model that suggests, chronic stressors including hormonal disparities, social stigma, and physical distress may generate secondary effects like sleep disturbances. Furthermore, academic and occupational stressors, poor sleep hygiene, and a lack of mental health literacy contribute to this outcome, particularly among working women [27].

Nevertheless, the prevalence of PCOS is not only a biological but also strongly embedded with psychological responses and socio-cultural pressures as suggested by the bio psychosocial framework [38]. In rural Bangladesh, where femininity is thoroughly linked to reproductive capability, irregular menstrual cycles and infertility frequently lead to increased stigmatization, social isolation, and internalized disgrace [39]. This study found the elevated levels of stress and anxiety observed among rural women with PCOS that internalize such expectations. Previous studies also showed the intense social pressure about body image and fertility, that make hinder their mental well-being [16]. These dynamics reflect the broader influence of symbolic interactionism that highlights how social interactions and cultural narratives shape individuals’ self-concepts, particularly when they depart from traditional social norms [40]. Considering the Rotterdam criteria, women who met two or three diagnostic criteria showed significantly higher prevalence rates for all four mental health disorders compared to those who met fewer or none of them. A previous study has shown similar findings, indicating that women diagnosed with PCOS according to the Rotterdam criteria demonstrate significantly higher levels of psychological distress [41,42].

In addition, this study highlights the crucial influence of residential location on mental health outcomes among women with PCOS, with those in rural areas experiencing more severe symptoms as well as higher levels of depression, anxiety, and insomnia compared to their urban counterparts. This is consistent with previous studies [43]. The structural strain theory asserts that such differences are result of the social structures and existing inequalities in the society due to lack of access to healthcare, education, and employment [38]. Moreover, rural health care systems typically lack the specific logistics required for dealing complex syndromes like PCOS.

The unequal circulation of medical substructure further leads to late interventions, intensifying both physical symptoms and psychological strain [44]. Consequently, women with PCOS in rural Bangladesh frequently face noteworthy psychological issues as a result of inadequate reproductive healthcare, delayed diagnosis, and insufficient mental health services, all of which exacerbate the condition’s mental health burden [45]. Afterwards, the present study also found that women in rural areas exhibited a higher propensity to fulfill multiple Rotterdam criteria, indicating a more pronounced clinical manifestation of PCOS. The findings correspond with previous studies, demonstrating that women with PCOS living in rural regions displayed more severe phenotypes than those in urban areas [46].

On the other hand, concern regarding body image is found as a critical psychological stress across all BMI categories [47]. This study showed that many women are dissatisfied with their appearance, specifically due to PCOS-related symptoms such as weight gain, acne, and hirsutism. These concerns were strongly related to depression and anxiety, consistent with previous findings [48]. The social pressure and body image framework also denotes that cultural standards of fineness and femaleness marginalize women whose appearance deviates from these norms [49].

Yet, relational dynamics has critical influence on the psychological well-being of women [50]. The study showed that women in romantic relationships were 2.767 times more probable to encounter depression than those who are single. It is also revealed in previous study that married women in Bangladesh scored above the cut-off point for depression [51]. Besides, the feminist sociological theory advocates that in patriarchal societies like Bangladesh, romantic and marital relationships often augment reproductive prospects, thereby endangering women to continual inspection over their fertility [52]. Furthermore, the labeling theory suggests that such women who are deviate from cultural norms surrounding fertility may be labeled as ‘defective’ or ‘incomplete,’ resulting in assumed social stigma as products of systemic gender inequality, which ultimately deteriorating mental health outcomes [53].

However, this study is not beyond limitations. There is a constraint of causal interpretations between PCOS and psychological outcomes since it is a cross-sectional design. Moreover, self-reported measures may present response bias, predominantly in conservative surroundings like rural Bangladesh, where talking about mental health and reproductive issues still perceived as taboo. Yet, further research can utilize longitudinal and triangulation approaches to discover the chronological associations between PCOS and mental health. Furthermore, investigating intersectional factors including gender norms, healthcare access, and relational dynamics, across varied populations will provide deeper perceptions into socially delicate interventions for PCOS-related mental distress.

## 5.0 Conclusion

Polycystic ovary syndrome is a clinically significant hormonal condition affecting women of reproductive age. The rising prevalence of mental health issues is becoming increasingly concerning for Bangladesh and globally. The findings indicated a greater prevalence of psychological factors in women diagnosed with PCOS, especially those living in rural areas. Besides, a significant association was identified between PCOS and elevated levels of all four mental health factors, namely depression, anxiety, stress, and insomnia, in reproductive-aged women in Bangladesh. Furthermore, women diagnosed with PCOS who satisfied two or more of the Rotterdam criteria demonstrated notably higher levels of psychological morbidities

In these circumstances, policymakers should implement interventions that include regular screening of psychological and physical wellbeing as integral components of PCOS treatment, particularly for rural women and those exhibiting the PCOS phenotype. Further, health ministry could plan some workshops on mental health to raise awareness and organize counseling sessions in rural parts in order to save them from such mental health problems and to use their capacity to its maximum potential. Future research should include intervention and longitudinal studies to enhance understanding of the causal pathways associated with PCOS and to develop interventions focused on improving the quality of life for women affected by this condition.

## Data Availability Statement

The data used to support the investigation’s conclusion is owned by the corresponding author.

